# The effect of coronary revascularization treatment timing on mortality in patients with stable ischemic heart disease in British Columbia

**DOI:** 10.1101/2024.04.24.24306272

**Authors:** Sean Hardiman, Guy Fradet, Lisa Kuramoto, Michael Law, Simon Robinson, Boris Sobolev

## Abstract

**Background:** Prior research has shown that patients with stable ischemic heart disease who undergo delayed coronary artery bypass graft (CABG) surgery face higher mortality rates than those who receive CABG within the time recommended by physicians. However, this research did not account for percutaneous coronary intervention (PCI), a widely available alternative to delayed CABG in many settings. We sought to establish whether there was a difference in mortality between timely PCI and delayed CABG.

**Methods:** We identified 25,520 patients 60 years or older who underwent first-time non-emergency revascularization for angiographically-proven, stable left main or multi-vessel ischemic heart disease in British Columbia between January 1, 2001, and December 31, 2016. We estimated unadjusted and adjusted mortality after index revascularization or last staged PCI for patients undergoing delayed CABG compared to timely PCI.

**Findings:** After adjustment with inverse probability of treatment weights, at three years, patients who underwent delayed CABG had a statistically significant lower mortality compared with patients who received timely PCI (4.3% delayed CABG, 13.5% timely PCI; risk ratio 0.32, 95% CI 0.24 – 0.40).

**Interpretation:** Patients who undergo CABG with delay have a lower risk of death than patients who undergo PCI within appropriate time. Our results suggest that patients who wish to receive CABG as their revascularization treatment will receive a mortality benefit over PCI as an alternative strategy.

## INTRODUCTION

In Canada, clinical need, resource allocation, and variation in demand determine how soon diagnosed coronary artery disease will be treated. Regional health authorities operate predominantly under a global budget funding model[1] that effectively caps the annual volume of procedures that a hospital can perform. Therefore, patients who require non-emergency revascularization by coronary artery bypass graft (CABG) surgery or percutaneous coronary intervention (PCI) may find their procedures are delayed during periods of higher demand for cardiac care or reduced supply of hospital services.[2] Access is further compromised during times of crisis, such as during the early waves of the COVID-19 pandemic throughout the country when non-emergency health care services were stopped and only emergency cases continued.[3,4]

Prior research has shown that patients waiting for CABG benefit from earlier timing of treatment.[5] Multiple randomized clinical trials in patients with stable multi-vessel disease and left main disease have refined indications for CABG and PCI. Patients with multi-vessel or left-main coronary artery disease who do not need emergency treatment should consider CABG rather than PCI[6], due to lower mortality in some populations, fewer post-procedural myocardial infarctions, and a reduced need for repeat revascularization. However, none of these trials included patients with substantial delays in CABG treatment and evidence shows that mortality after CABG worsens when the surgery is delayed.[5] Moreover, PCI is considered a reasonable alternative to CABG. Therefore, we established our research question: do the proportions of long-term mortality differ between patients with stable multi-vessel or left main ischemic heart disease who have delayed CABG compared to those who have timely PCI? In other words, what would happen if patients who could only have CABG delivered below standard instead had PCI delivered to standard?

## MATERIALS AND METHODS

This study follows the Strengthening the Reporting of Observational Studies in Epidemiology (STROBE) guidelines for the reporting of observational cohort studies.[7]

We conducted a cohort study of prospectively collected data amongst all patients in British Columbia (BC) who underwent isolated CABG surgery or PCI for the treatment of coronary artery disease. We obtained diagnostic catheterization, PCI, and isolated CABG records from the provincial registries maintained by Cardiac Services BC (CSBC), a program of the Provincial Health Services Authority (Vancouver, BC). CSBC is responsible for the planning, funding, and quality of specialized tertiary cardiac services in the province, including cardiac surgery and interventional cardiology services. We used CSBC’s diagnostic catheterization, CABG, and PCI registry data to establish a single record that represents an episode of care which contains all events occurring from diagnostic catheterization through to revascularization. We linked these care episodes to the BC Ministry of Health’s Discharge Abstract Database (DAD), which contains hospitalization records, and the BC Vital Statistics Deaths File, which contains deaths data. Finally, we linked this data set to Population Data BC’s Central Demographics File, which contains demographic data for all study participants. In BC, the five cardiac centres are overseen by CSBC. CSBC structures, including annual quality reviews, bring together surgeons and interventional cardiologists from across the province. Each cardiac centre operates using a heart team model, though implementation varies amongst sites.

The study consists of patients aged 60 years or older, who underwent non-emergency first-time revascularization for angiographically-proven, stable left main or multi-vessel ischemic heart disease in British Columbia, Canada, between January 1, 2001, and December 31, 2016 (Figure 1), criteria used in the ASCERT study.[8] We defined revascularization as either a PCI or an isolated CABG surgery. Patient age, extent of disease, and non-emergency status were identified using the Cardiac Services BC cardiac surgery and PCI registry data. Stable disease was identified using atherosclerotic heart disease code (ICD-10-CA I25.0, I25.1, I25.10; ICD-9 429.2 414.0) logged as type M (most responsible), type 1 (pre-admit comorbidity), type 2 (post-admit comorbidity), type 6 (proxy most responsible diagnosis), or types W, X, or Y (first, second, or third service transfers) in the DAD. The index event in this study is first-ever revascularization, by either PCI or CABG, within the study period of January 1, 2001, and December 31, 2016.

**Figure 1.**
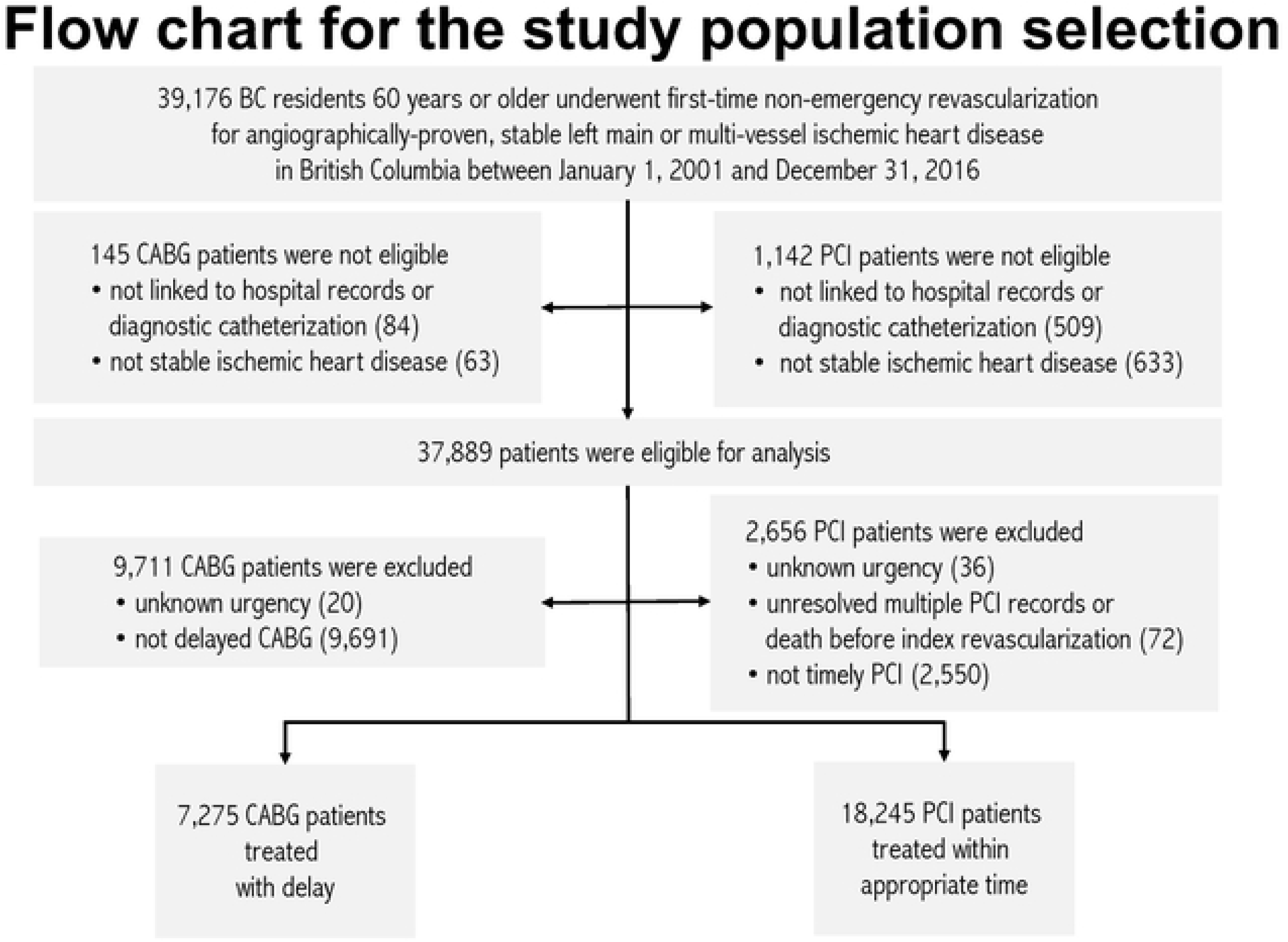
Flow chart for the study population.

### Variables

#### Study variable

The study variable is treatment timing, operationalized as the time to coronary revascularization treatment and computed in calendar days. Based on treatment timing and the type of revascularization procedure received, patients were assigned to one of two study groups: delayed CABG or timely PCI. The time to treatment starts on the date when the need for revascularization is clinically established and the patient is ready, willing, and able to undergo revascularization. The time to treatment ends on the date the index revascularization procedure was performed. To establish intervals defining timely and delayed treatment, we used the Canadian Cardiovascular Society (CCS) recommended times[9] to define delayed CABG and timely PCI for semi-urgent and elective CABG and PCI patients, the First Minsters’ Meeting benchmarks[10] for urgent CABG, and CSBC benchmarks for urgent PCI patients (Table 1).

**Table 1.** Study group assignments by procedure type and urgency and treatment delay in days.

| Procedure | Urgency | Timely Treatment |  | Delayed Treatment |
| --- | --- | --- | --- | --- |
|  |  | Interval Start | Interval End | Interval Start |
| CABG | Priority I | 1 Day | 7 Days | 8+ Days |
|  | Priority II | 1 Day | 14 Days | 15+ Days |
|  | Priority III | 1 Day | 42 Days | 43+ Days |
| PCI | Urgent Inpatient | 1 Day | 5 Days | 6+ Days |
|  | Urgent Outpatient | 1 Day | 14 Days | 15+ Days |
|  | Elective | 1 Day | 42 Days | 43+ Days |

Dates were collected from CSBC registries for CABG and PCI, where triage coordinators recorded the date that the patient was booked for their procedure and the date their procedure occurred.

#### Outcome variable

The outcome variable is the time to death in days from any cause recorded in the BC Vital Statistics Deaths File. We followed patients from the time of index revascularization or last staged PCI until death, the end of the study, or three years’ follow-up, whichever came first. Due to data limitations, we developed a rule (Supplementary Material) to differentiate staged PCI from repeat revascularization in patients with multiple PCI records, based on Spitzer’s[11] criteria (Supplementary Material).

#### Additional variables

We used variables in the form in which they were received from the data stewards. Some concepts, such as comorbidities and clearance time, were operationalized by the study team from data already in the data set (see Supplementary Material). Patients with a comorbidity identified as metastatic cancer process were grouped to the Metastatic Cancer variable. CABG patients whose CCS angina grade[12] was classified as Class 4A, 4B, or 4C in the CSBC cardiac surgery registry were grouped to Class 4.

### Statistical Methods

We estimated the frequency and percentage of patients by characteristics and by treatment group. Groups were compared using a chi-square test for categorical variables and p-values for between group differences reported. We modelled cumulative mortality over three years using with a flexible parametric approach. This approach uses restricted cubic spline functions, also known as Royston-Parmar models, to model the baseline mortality over three years.[13] The main advantage of this model is that it provides the means to smoothly estimate the survival function, in contrast to the Cox model, where the baseline hazard function can be a noisy step function.[13] We then estimated the unadjusted cumulative mortality proportion over three years for each study group. Finally, we estimated risk ratios comparing the treatment groups. in mortality at three years. A risk ratio of less than one means the delayed CABG group had a lower risk of mortality at 3 years compared to the timely PCI group. A risk ratio of greater than one means the delayed CABG group had a higher risk of mortality at 3 years compared to the timely PCI group. We selected three years’ follow-up as the follow-up time, considering the variation in follow-up time used in randomized controlled trials of CABG versus PCI.

We estimated propensity scores[14] for the probability of belonging to each study group using logistic regression and, using those scores, calculated inverse probability of treatment weights[15], following the ASCERT approach.[8] Inverse probability of treatment weighting creates a synthetic cohort that utilizes all patient information, compared to other propensity score methods where this cannot be assured. Variables were selected starting with those used in ASCERT[8] and informed by a scoping review of the factors of mortality after CABG.[16] Each patient was weighted by the inverse of the probability of being assigned to their treatment group to adjust for differences between the two treatment groups. We compared the performance of the propensity score model by comparing the distribution of covariates and propensity scores before and after inverse probability weighting. Adjusted mortality estimates were obtained using an inverse probability weighted flexible parametric approach. Statistical analyses were performed using Stata 17 (College Station, TX). Flexible parametric models were constructed using *stpm2*, a Stata software package.[17]

### Patient and public involvement

We consulted the Pacific Open-Heart Association (POHA) to inform development of our research question. POHA provides peer support to patients undergoing or who have undergone heart surgery in the Vancouver, BC area. They confirmed that BC patients often wait for CABG and that anxiety results when it isn’t known when a planned CABG will occur. In this study, we address the question that patients told us matters most: would it be better for CABG candidates to undergo PCI instead of facing an indeterminate length of time waiting for CABG?

### Data access

The authors first gained access to the data for research purposes on June 7, 2019, and had access to the data through April 30, 2025. The authors had no access to information that could identify individual participants during or after data collection.

## RESULTS

### Setting and Participants

We identified 39,176 British Columbia patients who met the selection criteria for our study (Figure 1).

We did not select patients for the analytical cohort if their revascularization record could not be linked to hospital records or their PCI record was for ad-hoc PCI, but the procedure could not be linked to a diagnostic catheterization (*n*=591), or that their hospital records did not contain diagnosis codes indicative of stable ischemic heart disease (*n*=696). 37,889 patients were eligible for analysis. We set aside patients if their procedure urgency could not be determined (*n*=56), if patients with multiple PCI records were unresolved after applying the repeat revascularization algorithm or if there were errors in the administrative data set where date of death preceded date of revascularization (*n*=72), if the patient received delayed PCI (*n*=2,550), or if the patient received timely CABG (*n*=9,711). 25,520 patients were available to be analyzed.

### Descriptive data

Table 2 shows the baseline characteristics of patients in the study cohort.

**Table 2.** Baseline characteristics of the patients.

|  | Unadjusted Data |  |  |  |  | Data Adjusted with<br>Inverse Probability Weighting |  |  |  |  |
| --- | --- | --- | --- | --- | --- | --- | --- | --- | --- | --- |
|  | Timely PCI<br>(n= 18,245) |  | Delayed<br>CABG<br>(n= 7,275) |  | P-Value | Timely PCI<br>(n= 26,376) |  | Delayed CABG<br>(n= 22,813) |  | P-Value |
|  | N | % | N | % |  | N | % | N | % |  |
| Age* |  |  |  |  |  |  |  |  |  |  |
| 60 - 64 | 3,594 | 19.7% | 1,609 | 22.1% | <0.00 | 5,150 | 19.5% | 4,556 | 20.0% | 0.17 |
| 65 - 69 | 3,849 | 21.1% | 1,868 | 25.7% |  | 5,862 | 22.2% | 4,760 | 20.9% |  |
| 70 - 74 | 3,635 | 19.9% | 1,827 | 25.1% |  | 5,811 | 22.0% | 5,950 | 26.1% |  |
| 75 - 79 | 3,361 | 18.4% | 1,355 | 18.6% |  | 4,791 | 18.2% | 4,225 | 18.5% |  |
| >=80 | 3,806 | 20.9% | 616 | 8.5% |  | 4,762 | 18.1% | 3,322 | 14.6% |  |
| Sex |  |  |  |  |  |  |  |  |  |  |
| Male | 12,718 | 69.7% | 5,994 | 82.4% | <0.00 | 19,389 | 73.5% | 16,323 | 71.6% | 0.49 |
| Female | 5,527 | 30.3% | 1,281 | 17.6% |  | 6,987 | 26.4% | 6,489 | 28.4% |  |
| Body Mass Index* |  |  |  |  |  |  |  |  |  |  |
| <18.5 | 237 | 1.3% | 43 | 0.6% | <0.00 | 298 | 1.1% | 130 | 0.6% | 0.52 |
| ≥18.5 and <25 | 5,380 | 29.5% | 1,897 | 26.1% |  | 7,583 | 28.8% | 6,718 | 29.4% |  |
| ≥25 and <30 | 7,886 | 43.2% | 3,173 | 43.6% |  | 11,452 | 43.4% | 9,983 | 43.8% |  |
| >30 | 4,625 | 25.3% | 2,046 | 28.1% |  | 6,857 | 26.0% | 5,777 | 25.3% |  |
| Missing | 117 | 0.6% | 116 | 1.6% |  | 186 | 0.7% | 205 | 0.9% |  |
| Extent of Disease |  |  |  |  |  |  |  |  |  |  |
| Double Vessel Disease | 10,076 | 55.2% | 543 | 7.5% | <0.00 | 10,575 | 40.1% | 8,350 | 36.6% | 0.18 |
| Triple Vessel Disease | 7,290 | 40.0% | 4,587 | 63.1% |  | 11,643 | 44.1% | 11,151 | 48.9% |  |
| Left Main Disease | 879 | 4.8% | 2,145 | 29.5% |  | 4,158 | 15.8% | 3,311 | 14.5% |  |
| Ejection Fraction† |  |  |  |  |  |  |  |  |  |  |
| EF <30% | 697 | 3.8% | 268 | 3.7% | <0.00 | 984 | 3.7% | 830 | 3.6% | 0.70 |
| EF ≥30% and ≤50% | 3,389 | 18.6% | 1,965 | 27.0% |  | 5,765 | 21.9% | 4,550 | 19.9% |  |
| EF >50% | 10,410 | 57.1% | 4,496 | 61.8% |  | 15,244 | 57.8% | 13,385 | 58.7% |  |
| Missing | 3,749 | 20.5% | 546 | 7.5% |  | 4,383 | 16.6% | 4,047 | 17.7% |  |
| Serum Creatinine (μmol/L)* |  |  |  |  |  |  |  |  |  |  |
| <60 | 775 | 4.2% | 219 | 3.0% | <0.00 | 977 | 3.7% | 1,281 | 5.6% | 0.47 |
| 60≥ and <80 | 4,380 | 24.0% | 1,525 | 21.0% |  | 6,149 | 23.3% | 5,308 | 23.3% |  |
| 80≥ and <99 | 6,400 | 35.1% | 2,470 | 34.0% |  | 9,332 | 35.4% | 8,134 | 35.7% |  |
| ≥100 | 6,024 | 33.0% | 2,320 | 31.9% |  | 8,562 | 32.5% | 6,813 | 29.9% |  |
| Unknown | 666 | 3.7% | 741 | 10.2% |  | 1,355 | 5.1% | 1,276 | 5.6% |  |
| Canadian Cardiovascular Society Angina Class* |  |  |  |  |  |  |  |  |  |  |
| None | 715 | 3.9% | 389 | 5.3% | <0.00 | 1,082 | 4.1% | 1,161 | 5.1% | 0.12 |
| Class 1 | 661 | 3.6% | 347 | 4.8% |  | 970 | 3.7% | 1,020 | 4.5% |  |
| Class 2 | 2,882 | 15.8% | 1,785 | 24.5% |  | 4,872 | 18.5% | 4,473 | 19.6% |  |
| Class 3 | 1,452 | 8.0% | 2,730 | 37.5% |  | 4,724 | 17.9% | 4,030 | 17.7% |  |
| Class 4 | 11,450 | 62.8% | 1,494 | 20.5% |  | 12,821 | 48.6% | 10,113 | 44.3% |  |
| Atypical | 314 | 1.7% | 68 | 0.9% |  | 377 | 1.4% | 208 | 0.9% |  |
| Missing | 771 | 4.2% | 462 | 6.4% |  | 1,529 | 5.8% | 1,807 | 7.9% |  |
| Prior Acute Myocardial Infarction* |  |  |  |  |  |  |  |  |  |  |
| Yes | 3,479 | 19.1% | 2,516 | 34.6% | <0.00 | 6,843 | 25.9% | 5,993 | 26.3% | 0.98 |
| Unknown | 4,385 | 24.0% | 2,109 | 29.0% |  | 6,616 | 25.1% | 5,654 | 24.8% |  |
| Smoking Status* |  |  |  |  |  |  |  |  |  |  |
| Never | 7,094 | 38.9% | 2,626 | 36.1% | <0.00 | 9,620 | 36.5% | 8,798 | 38.6% | 0.34 |
| Current/Now | 2,283 | 12.5% | 621 | 8.5% |  | 2,979 | 11.3% | 2,181 | 9.6% |  |
| Former/Quit | 8,226 | 45.1% | 3,086 | 42.4% |  | 12,129 | 46.0% | 10,054 | 44.1% |  |
| Unknown | 642 | 3.5% | 942 | 12.9% |  | 1,648 | 6.2% | 1,780 | 7.8% |  |
| Comorbidities |  |  |  |  |  |  |  |  |  |  |
| Atrial Fibrillation or Atrial Flutter | 1,421 | 7.8% | 2,051 | 28.2% | <0.00 | 3,210 | 12.2% | 3,236 | 14.2% | 0.14 |
| Cardiac Dysrhythmias§ | 728 | 4.0% | 351 | 4.8% | 0.00 | 1,083 | 4.1% | 1,477 | 6.5% | 0.05 |
| Cerebrovascular Disease | 428 | 2.3% | 379 | 5.2% | <0.00 | 906 | 3.4% | 1,251 | 5.5% | 0.27 |
| Chronic Pulmonary Disease | 911 | 5.0% | 341 | 4.7% | 0.31 | 1,465 | 5.6% | 982 | 4.3% | 0.23 |
| Congestive Heart Failure | 2,109 | 11.6% | 958 | 13.2% | <0.00 | 3,474 | 13.2% | 2,496 | 10.9% | 0.09 |
| Connective Tissue Disease | 238 | 1.3% | 93 | 1.3% | 0.87 | 356 | 1.3% | 280 | 1.2% | 0.78 |
| Diabetes | 4,986 | 27.3% | 2,908 | 40.0% | <0.00 | 8,484 | 32.2% | 7,416 | 32.5% | 0.88 |
| Hypertension | 9,102 | 49.9% | 4,450 | 61.2% | <0.00 | 14,237 | 54.0% | 11,814 | 51.8% | 0.39 |
| Hypertensive Heart Disease | 35 | 0.2% | 18 | 0.2% | 0.38 | 149 | 0.6% | 48 | 0.2% | 0.21 |
| Liver Disease | 53 | 0.3% | 19 | 0.3% | 0.69 | 105 | 0.4% | 72 | 0.3% | 0.67 |
| Metastatic Cancer | 496 | 2.7% | 168 | 2.3% | 0.06 | 662 | 2.5% | 662 | 2.9% | 0.67 |
| Peripheral Vascular Disease | 715 | 3.9% | 460 | 6.3% | <0.00 | 1,404 | 5.3% | 1,064 | 4.7% | 0.55 |
| Pneumonia | 522 | 2.9% | 287 | 3.9% | <0.00 | 1,088 | 4.1% | 776 | 3.4% | 0.50 |
| Renal Disease | 1,240 | 6.8% | 711 | 9.8% | <0.00 | 1,895 | 7.2% | 1,770 | 7.8% | 0.59 |
| Ulcer Disease | 104 | 0.6% | 104 | 1.4% | <0.00 | 179 | 0.7% | 494 | 2.2% | 0.01 |
| Calendar Period of Index Revascularization |  |  |  |  |  |  |  |  |  |  |
| 2001 | 949 | 5.2% | 484 | 6.7% | <0.00 | 1,532 | 5.8% | 1,901 | 8.3% | 0.26 |
| 2002 | 1,101 | 6.0% | 684 | 9.4% |  | 1,766 | 6.7% | 2,088 | 9.2% |  |
| 2003 | 1,210 | 6.6% | 714 | 9.8% |  | 2,023 | 7.7% | 2,180 | 9.6% |  |
| 2004 | 1,224 | 6.7% | 544 | 7.5% |  | 1,826 | 6.9% | 1,612 | 7.1% |  |
| 2005 | 1,166 | 6.4% | 503 | 6.9% |  | 1,844 | 7.0% | 1,499 | 6.6% |  |
| 2006 | 1,100 | 6.0% | 512 | 7.0% |  | 1,688 | 6.4% | 1,260 | 5.5% |  |
| 2007 | 1,252 | 6.9% | 485 | 6.7% |  | 1,690 | 6.4% | 1,353 | 5.9% |  |
| 2008 | 1,266 | 6.9% | 373 | 5.1% |  | 1,797 | 6.8% | 1,256 | 5.5% |  |
| 2009 | 1,342 | 7.4% | 238 | 3.3% |  | 1,790 | 6.8% | 1,592 | 7.0% |  |
| 2010 | 1,387 | 7.6% | 263 | 3.6% |  | 1,639 | 6.2% | 945 | 4.1% |  |
| 2011 | 1,325 | 7.3% | 250 | 3.4% |  | 1,576 | 6.0% | 1,084 | 4.8% |  |
| 2012 | 1,122 | 6.1% | 347 | 4.8% |  | 1,467 | 5.6% | 1,094 | 4.8% |  |
| 2013 | 897 | 4.9% | 467 | 6.4% |  | 1,470 | 5.6% | 1,373 | 6.0% |  |
| 2014 | 939 | 5.1% | 455 | 6.3% |  | 1,434 | 5.4% | 1,295 | 5.7% |  |
| 2015 | 905 | 5.0% | 421 | 5.8% |  | 1,296 | 4.9% | 940 | 4.1% |  |
| 2016 | 1,060 | 5.8% | 535 | 7.4% |  | 1,537 | 5.8% | 1,339 | 5.9% |  |
| Hospital Type |  |  |  |  |  |  |  |  |  |  |
| Metropolitan | 14,197 | 77.8% | 5,396 | 74.2% | <0.00 | 20,531 | 77.8% | 16,981 | 74.4% | 0.17 |
| Urban | 4,048 | 22.2% | 1,879 | 25.8% |  | 5,845 | 22.2% | 5,831 | 25.6% |  |
| Clearance Time Category¶ |  |  |  |  |  |  |  |  |  |  |
| 1 Week | 14,410 | 79.0% | 3,052 | 42.0% | <0.00 | 17,410 | 66.0% | 14,689 | 64.4% | 0.66 |
| 2 Weeks | 2,387 | 13.1% | 1,910 | 26.3% |  | 4,836 | 18.3% | 4,332 | 19.0% |  |
| 3 or More Weeks | 1,448 | 7.9% | 2,313 | 31.8% |  | 4,130 | 15.7% | 3,791 | 16.6% |  |

| Neighborhood Income Decile |  |  |  |  |  |  |  |  |  |  |
| --- | --- | --- | --- | --- | --- | --- | --- | --- | --- | --- |
| Lowest Decile | 1,956 | 10.7% | 707 | 9.7% | 0.22 | 2,664 | 10.1% | 2,417 | 10.6% | 0.87 |
| 2nd Decile | 1,925 | 10.6% | 756 | 10.4% |  | 2,814 | 10.7% | 2,089 | 9.2% |  |
| 3rd Decile | 1,822 | 10.0% | 742 | 10.2% |  | 2,742 | 10.4% | 2,427 | 10.6% |  |
| 4th Decile | 1,907 | 10.5% | 784 | 10.8% |  | 2,934 | 11.1% | 2,790 | 12.2% |  |
| 5th Decile | 1,778 | 9.7% | 759 | 10.4% |  | 2,374 | 9.0% | 2,030 | 8.9% |  |
| 6th Decile | 1,760 | 9.6% | 645 | 8.9% |  | 2,392 | 9.1% | 2,249 | 9.9% |  |
| 7th Decile | 1,699 | 9.3% | 694 | 9.5% |  | 2,459 | 9.3% | 2,097 | 9.2% |  |
| 8th Decile | 1,755 | 9.6% | 708 | 9.7% |  | 2,774 | 10.5% | 2,131 | 9.3% |  |
| 9th Decile | 1,698 | 9.3% | 706 | 9.7% |  | 2,421 | 9.2% | 1,957 | 8.6% |  |
| Highest Decile | 1,679 | 9.2% | 673 | 9.3% |  | 2,346 | 8.9% | 2,360 | 10.3% |  |
| Unknown | 266 | 1.5% | 101 | 1.4% |  | 457 | 1.7% | 265 | 1.2% |  |

\* At the time of revascularization.
† Ejection Fraction at the time of revascularization; if missing, at the time of diagnostic catheterization.
§ Excluding atrial fibrillation and atrial flutter.
¶ Clearance time is the hypothetical time within which the wait list would be cleared at maximum weekly service capacity if there were no new arrivals.

Before adjustment with inverse probability weighting, the patients undergoing delayed CABG were, compared to patients undergoing timely PCI, had higher proportions of male sex, a BMI >30, triple vessel disease, left main disease, and an ejection fraction ≤50%. The delayed CABG group had significantly higher proportions of atrial fibrillation or atrial flutter, congestive heart failure, diabetes, hypertension, and renal disease, compared to timely PCI. The timely PCI group had higher proportions of double-vessel disease and Canadian Cardiovascular Society (CCS) Angina Class 4. Most patients, regardless of study group, were treated in metropolitan hospitals. Clearance time is shorter amongst patients treated with timely PCI compared to delayed CABG. Proportions of neighborhood income decile are balanced throughout the study cohort. Of the patients who underwent PCI, 48.1% received Bare-Metal Stents (BMS), 4.5% received a combination of BMS and Drug-Eluting Stents (DES) and 42.8% received only DES. Of the patients who underwent CABG, 8.5% received only a saphenous vein graft, 71.6% received a single arterial graft, 16.3% received a double arterial graft, and 3.4% received a triple arterial graft. The mean waiting time for the timely PCI group was 11.9 days and the mean waiting time for the delayed CABG group was 71.5 days (Supplementary Material).

As expected, patients in the timely PCI group had a lower probability of being selected for delayed CABG than did those in the CABG group. However, all patients had a positive probability of being assigned to either CABG or PCI, consistent with results of comparative effectiveness studies of CABG and PCI published elsewhere.[8]

### Outcome data and main results

Unadjusted failure curves are shown in Figure 2; unadjusted cumulative mortality and risk ratios are shown in Table 3.

**Figure 2.**
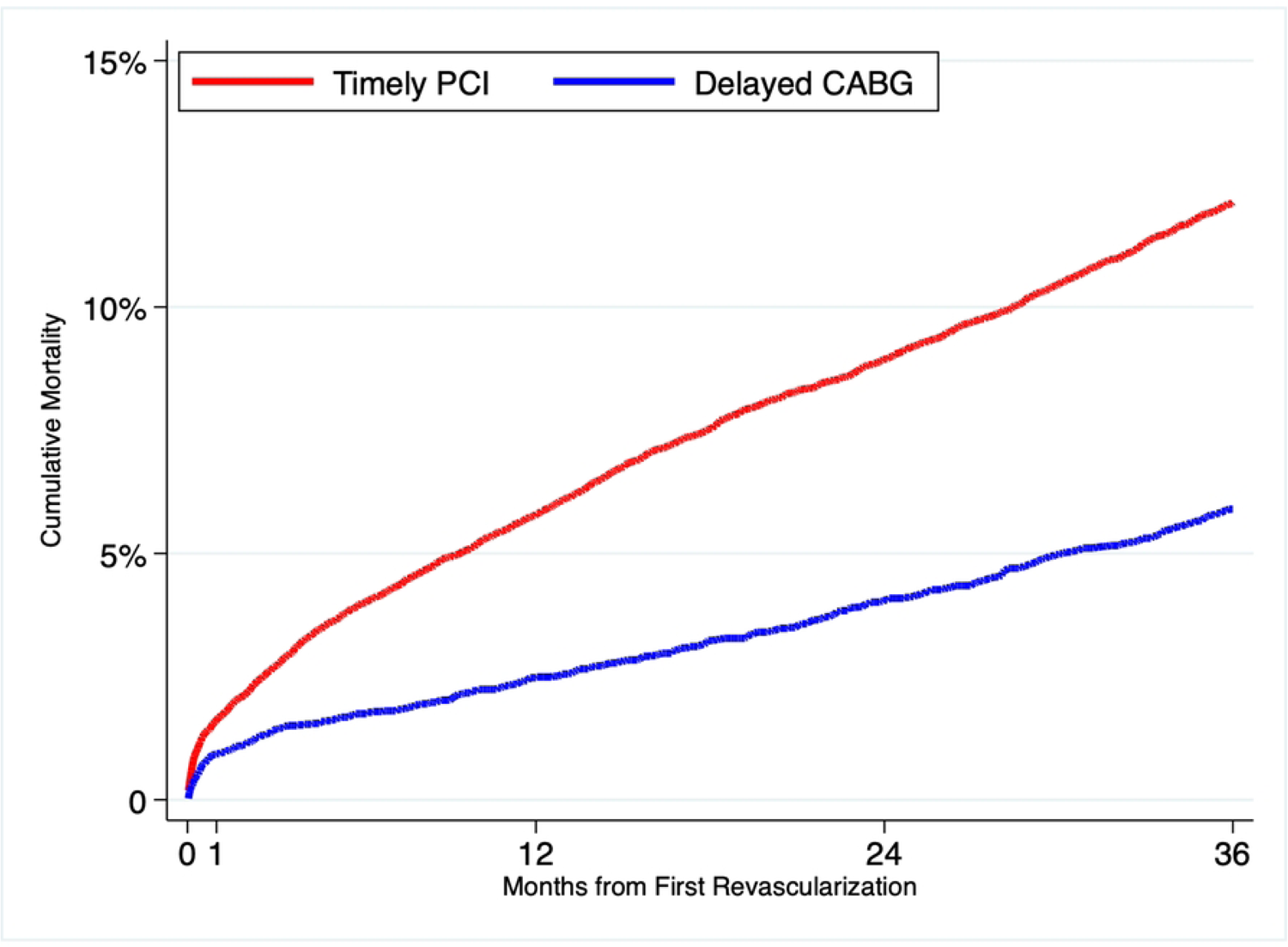
Cumulative mortality in the CABG and PCI populations, from an unadjusted analysis.

**Table 3.** Rates of mortality (percent), risk ratios, and 95% confidence intervals in the delayed CABG and timely PCI populations, from an unadjusted analysis.

|  | **30 Days** | **1 Year** | **2 Years** | **3 Years** |
| --- | --- | --- | --- | --- |
| Delayed CABG | 0.7 (0.6, 0.9) | 2.6 (2.3, 2.9) | 4.2 (3.8, 4.6) | 5.9 (5.3, 6.4) |
| Timely PCI | 1.5 (1.4, 1.7) | 5.8 (5.4, 6.1) | 8.9 (8.5, 9.3) | 12.1 (11.6, 12.6) |
| Risk Ratio for Delayed CABG | 0.48 (0.36, 0.60) | 0.45 (0.39, 0.52) | 0.47 (0.42, 0.52) | 0.48 (0.43, 0.53) |

At 30 days, there was a significant difference in unadjusted mortality between the groups (0.7% in the delayed CABG group compared with 1.5% in the timely PCI group). The 1-year unadjusted mortality rate was 2.6% in the delayed CABG group and 5.8% in the timely PCI group. The 2-year unadjusted mortality rate was 4.2% in the delayed CABG group and 8.9% in the timely PCI group. The 3-year unadjusted mortality rate was 5.9% in the delayed CABG group and 12.1% in the timely PCI group (risk ratio [RR] 0.48, 95% confidence interval [CI], 0.43 – 0.53).

Failure curves adjusted with inverse probability of treatment weighting are shown in Figure 3; adjusted cumulative mortality and risk ratios are shown in Table 4.

**Figure 3.**
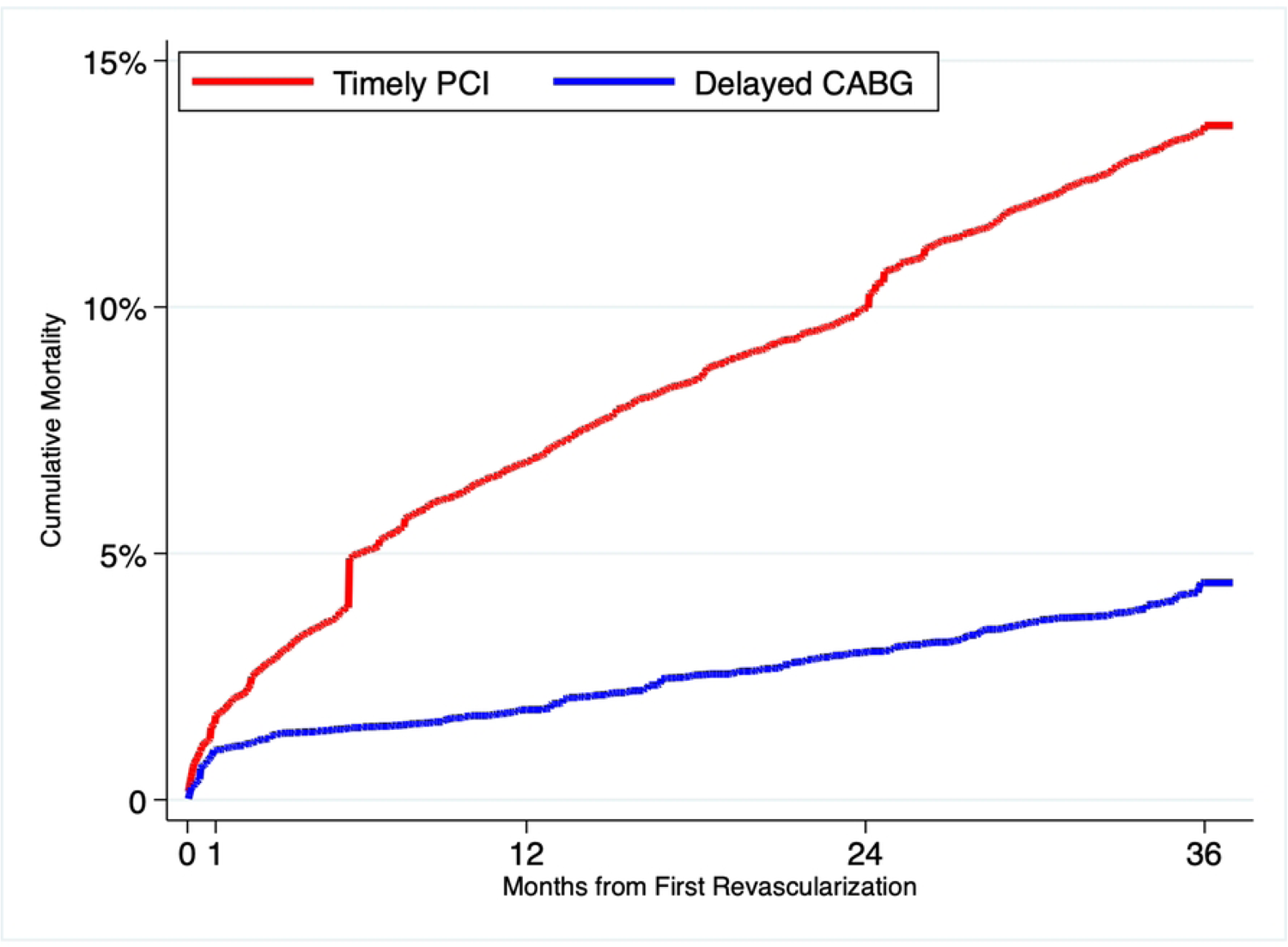
Cumulative mortality in the CABG and PCI populations, from an analysis adjusted with the use of inverse probability of treatment weighting.

**Table 4.**
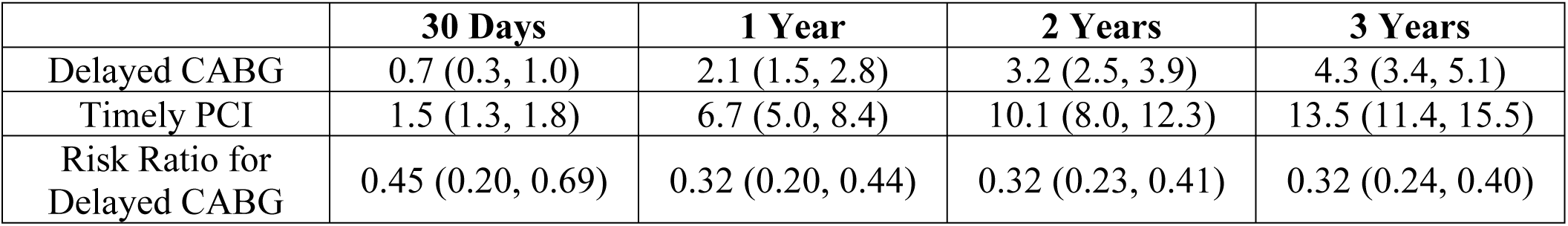
Rates of mortality (percent), risk ratios, and 95% confidence intervals in the delayed CABG and timely PCI populations, from an adjusted analysis.

|  | **30 Days** | **1 Year** | **2 Years** | **3 Years** |
| --- | --- | --- | --- | --- |
| Delayed CABG | 0.7 (0.3, 1.0) | 2.1 (1.5, 2.8) | 3.2 (2.5, 3.9) | 4.3 (3.4, 5.1) |
| Timely PCI | 1.5 (1.3, 1.8) | 6.7 (5.0, 8.4) | 10.1 (8.0, 12.3) | 13.5 (11.4, 15.5) |
| Risk Ratio for Delayed CABG | 0.45 (0.20, 0.69) | 0.32 (0.20, 0.44) | 0.32 (0.23, 0.41) | 0.32 (0.24, 0.40) |

At 30 days, there was a significant difference in adjusted mortality between the groups (0.7% in the delayed CABG group compared with 1.5% in the timely PCI group, RR 0.32; 95% CI 0.24 - 0.40). The 1-year adjusted mortality was 2.1% in the delayed CABG group and 6.7% in the timely PCI group. The 2-year adjusted mortality was 3.2% in the delayed CABG group compared with 10.1% in the timely PCI group. The 3-year adjusted mortality rate was 4.3% in the delayed CABG group and 13.5% in the timely PCI group (risk ratio 0.32, 95% CI, 0.24 to 0.40).

## DISCUSSION

### Key results

This study used data from multiple population-based registries and databases to evaluate the effectiveness of delayed CABG as compared with timely PCI. In this study, we found that amongst British Columbia patients 60 aged years or older, who underwent non-emergency first-time revascularization for angiographically-proven, stable left main or multi-vessel ischemic heart disease in British Columbia, between January 1, 2001, and December 31, 2016, there was a significant difference in both unadjusted mortality and mortality adjusted using inverse probability of treatment weights at 30 days, one year, two years, and three years. When the study cohort was stratified into three time periods, results were consistent with the results observed in the primary analysis (Supplementary Material).

Our findings should be considered in the context of results from other studies. There have been seven randomized, controlled trials comparing CABG with balloon angioplasty, eleven comparing CABG with PCI and stenting in patients with multi-vessel disease and six comparing CABG with PCI and stenting in specifically in patients with left-main disease. A survival advantage for CABG was noted in the Stent or Surgery (SoS) trial at two years [18] and sustained at six years.[19] In the ARTS-I trial, where BMS were used, a survival advantage at five years was not found.[20] However, the ARTS-II trial, which compared the ARTS-I CABG cohort to a new PCI cohort where patients received DES, did find a statistically significant difference in mortality. The FREEDOM trial, where diabetics with multivessel disease were treated with CABG compared to PCI with DES also found a statistically significant difference in mortality at five years,[21] and again at seven years,[22] consistent with the subgroup analysis reported by the BARI Investigators,[23] but inconsistent with the SYNTAX diabetes subgroup study that showed no statistical difference,[24] though a trend to better outcomes with CABG was observed. In the SYNTAX study, no difference in all-cause mortality was observed at three years, however, a statistically significant benefit was found for CABG patients with triple-vessel disease.[25] The benefits for patients with triple-vessel disease were sustained in the ten-year SYNTAX extension study.[26]

Of note is the finding of a difference in mortality at 30 days of index revascularization or last staged PCI. Randomized controlled trials and observational studies have shown higher mortality in patients undergoing CABG during the first 30 days compared to patients undergoing PCI, attributed to increased risk of death in the immediate post-operative period. We selected only patients with stable ischemic heart disease, who would be expected to have lower surgical risk than patients with more unstable disease. This may account for lower early mortality observed in this study. BC also has a long-standing cardiac surgery quality oversight program, delivered by CSBC in collaboration with the cardiac surgery community. Annual reporting on hospital and surgeon mortality at 30 days and 1 year during the study period may have contributed to improvements in the structures and processes associated with care quality that could contribute to lower mortality.

Observational studies also inform our understanding of these results. The ASCERT study[8] examined the comparative effectiveness of CABG versus PCI in Medicare patients using patient selection criteria and analytical methods common to ours. While no significant difference in mortality was found after one year, lower mortality was observed with CABG compared to PCI at four years, similar to our results. Similar findings were recently reported by Mehaffey and colleagues using contemporary CABG and PCI techniques in Medicare patients.[27] Our findings are consistent with those of a recent systematic review[28] included 23 studies comparing CABG with PCI. CABG was associated with better survival during follow-up in 17 studies and no significant difference between treatments in six, with no study favouring PCI. A recent meta-analysis reported similar results in patients who received CABG compared to PCI with DES.[29]

These results are noteworthy in that they demonstrate that amongst this patient population, the benefits of CABG do not appear to be attenuated by delay compared to PCI. For physicians who must advise patients on treatment strategies in the context of scarce resources, these results suggest that PCI as an alternative revascularization strategy may not be indicated if a reduced risk of mortality is desired. Patients can know that waiting for CABG may have benefits over PCI. Policymakers should interpret these results in the context of past CABG research, which shows benefit to earlier timing of treatment.[5]

### Limitations

This study has several limitations. First, we used inverse probability of treatment weighting to balance differences in patient and system factors between our study groups. While balance was achieved between our study groups, it is possible unmeasured confounding affected our findings. Second, we studied patients who underwent treatment between 2001 and 2016, during which stent technology evolved significantly, and the use of antiplatelet therapy evolved significantly. While we accounted for this by adjusting for calendar year of procedure, this may not have been sufficient to address the effect of time on outcomes. Third, we were limited by data available from CSBC in how we could establish extent of disease. While revascularization appropriate use criteria[30] suggest the use of SYNTAX scores to differentiate eligibility for CABG or PCI, this data is not routinely collected in BC. While SYNTAX scores are thought to have limited utility due to inter-rater variability,[6] the absence of this data limited our ability to stratify our patient groups to match those proposed in appropriate use criteria.[30] Finally, clinical presentation data was not consistently available during the study period, so we used diagnosis codes to identify patients with stable disease and did not select any patient with an ‘emergency’ priority for our study cohort. These efforts may not have completely excluded patients with more serious acuity who were not eligible for CABG and instead treated with PCI.

### Interpretation

Our results suggest that there is evidence that the treatment benefit of CABG surgery is not attenuated because of a delay in treatment when compared to PCI provided within appropriate time.

### Generalizability

As a population-based study, our results can be generalized to similar populations as those selected for this study. Our results can also be generalized to cardiac services systems with structures and processes that align with those that found in BC. Caution should be taken in applying these results to other populations and systems.

## CONCLUSION

In summary, this study used data from the CSBC diagnostic catheterization, PCI, and CABG registries, linking to the DAD, the BC Vital Statistics Deaths File and Population Health Data BC’s Central Demographics File to assess the comparative effectiveness of timely PCI and delayed CABG. We found that amongst patients older than 60 years of age with stable, multi-vessel or left-main ischemic heart disease that did not require emergency treatment that there was a statistically significant short-term and long-term survival advantage for patients who underwent delayed CABG compared to those who had timely PCI. Patients who face extended waiting times for CABG should be aware of these benefits before choosing PCI as an alternative revascularization strategy. Given these findings and the continued evolution of both CABG and PCI procedures, further research on the effects of delay is indicated.

## Data Availability

Access to data provided by the Data Stewards is subject to approval but can be requested for research projects through the Data Stewards or their designated service providers. All inferences, opinions, and conclusions drawn in this publication are those of the author(s), and do not reflect the opinions or policies of the Data Stewards.

## CONTRIBUTORS

BS and GF conceived the original study idea. SH, SB, ML, SR, and GF contributed to acquiring the financial support for the study. SH designed the study and acquired the data from the data stewards. SH designed the dataset creation plan (DCP) and the plan of analysis (POA). SH and LK created the analytical data set. SH performed all statistical analyses in the plan of analysis; LK performed statistical code review. All authors contributed to the content of the DCP and POA. All authors contributed to the interpretation of the analyzed data. SH wrote the original draft of the manuscript. All authors critically revised the manuscript. All authors approved the final version for submission. BS supervised the research project.

## ETHICS APPROVAL

The University of British Columbia Clinical Research Ethics Board (Certificate H17-00505) provided ethical approval for this research.

## PATIENT CONSENT STATEMENT

The authors confirm that patient consent is not applicable to this article. This is a retrospective analysis of de-identified data stored in registries and databases. Thus, the University of British Columbia Clinical Research Ethics Board did not require consent from patients to conduct this study.

## LIST OF ABBREVIATIONS

BC: British Columbia
BMS: Bare-Metal Stent
CABG: Coronary Artery Bypass Graft
CCS: Canadian Cardiovascular Society
CSBC: Cardiac Services BC
DAD: Discharge Abstract Database
DES: Drug-Eluting Stent
PCI: Percutaneous Coronary Intervention
POHA: Pacific Open Heart Association
STROBE: Strengthening the Reporting of Observational Studies in Epidemiology

